# Deep Learning Fusion for COVID-19 Diagnosis

**DOI:** 10.1101/2020.12.11.20246546

**Authors:** Odysseas Kechagias-Stamatis, Nabil Aouf, John A. Koukos

## Abstract

The outbreak of the novel coronavirus (COVID-19) disease has spurred a tremendous research boost aiming at controlling it. Under this scope, deep learning techniques have received even more attention as an asset to automatically detect patients infected by COVID-19 and reduce the doctor’s burden to manually assess medical imagery. Thus, this work considers a deep learning architecture that fuses the layers of current-state-of-the-art deep networks to produce a new structure-fused deep network. The advantages of our deep network fusion scheme are multifold, and ultimately afford an appealing COVID-19 automatic diagnosis that outbalances current deep learning methods. Indeed, evaluation on Computer Tomography (CT) and X-ray imagery considering a two-class (COVID-19/ non-COVID-19) and a four-class (COVID-19/ non-COVID-19/ Pneumonia bacterial / Pneumonia virus) classification problem, highlights the classification capabilities of our method attaining 99.3% and 100%, respectively.

## I. Introduction

Coronavirus Disease 2019 (COVID-19) is a respiratory syndrome affecting people on the entire globe and therefore it has been upgraded to a pandemic [1]. The number of infected is rapidly increasing on a daily basis posing a requirement for accurate and rapid diagnosis of COVID-19 patients to quarantine the suspect cases and deter the virus spread. To this end, COVID-19 diagnosis is mainly based on the Reverse Transcript Polymerase Chain Reaction (RT-PCR) [2]. However, the limited supply, requirements for laboratory environment, and high false-negative rates [3] affect the timely and accurate diagnosis of suspected patients, posing a mediocre prevention towards the spread of the infection. Thus, medical imagery and specifically Computerized Tomography (CT) and X-ray imagery are also exploited, either for COVID-19 infection cross-check or to speed-up the diagnosis process. Though, assessing medical imaging is currently a manual and time-consuming procedure imposing delays to the infection diagnosis process.

Hence, spurred by the recent advances of deep learning in various domains ranging from object classification [4], [5] to odometry [6], several automatic COVID-19 diagnosis methods have been proposed that exploit CT or X-ray imagery [7], [8]. Current techniques may utilize existing pre-trained deep learning models combined with transfer learning [9], [10], or use custom networks [11]–[13]. Pre-trained models are mostly trained on the ImageNet dataset [14] that despite including images from the visual domain, i.e. objects, animals, etc., partially re-training and fine-tuning these pre-trained models on medical imagery via transfer learning [15], adjusts the weights and bias of the model to accurately classify medical imagery. Typical pre-trained models used for COVID-19 diagnosis are AlexNet [16], GoogleNet [17], Visual Geometry Group (VGG) [18], ResNet [19] and inception [20]. Custom networks are also widely used and require being fully trained from scratch, i.e. the model weights and bias are the initialization values and need to be fully configured during the training process. However, the majority of these models are inspired by current pre-trained models that are properly tailored to meet the specific requirements of medical imagery classification.

Despite the ability of current deep learning models to diagnose COVID-19 patients, the classification power of these methods is limited by the deep learning structure itself. This is because each pre-trained or custom model presents its own novelties but also its own limitations that originate from its network structure and layer types. Hence, spurred by this finding, we propose a deep learning fusion scheme that interconnects the inner layers of each contributing deep network, i.e. sub-network, enhancing the classification strength of the fused network and minimizing its deficiencies. This is because during transfer learning the weights and bias of the formerly distinct networks are now cross-tuned and thus each sub-network affects the training process of its counterpart sub-network. To the best of our knowledge, this is a novel concept, both in the medical and in the computer vision domain.

The contributions of our paper can be summarized to:

a. Innovatively fusing current state-of-the-art pre-trained deep networks by cross-connecting their internal layers.
b. Creating deep and parallel networks for enhanced classification performance without the requirement of redesigning them.
c. Exploiting the knowledge encapsulated in each pre-trained network via transfer learning/ fine-tuning their weights within the fused network architecture. During transfer learning, the weights and bias of each sub-network are not only tuned based on the input imagery but are also affected by the counterpart sub-network.
d. The proposed deep learning scheme utilizes the advantages of each contributing deep network, i.e. residual module, inception module, etc., but still partially preserves its original capabilities as the sub-network structure is still preserved.
e. Despite the fused network being deep, it still preserves a low number of parameters compared to state-of-the-art deep networks of similar depth. This is important as fewer parameters contribute to faster training.

The rest of the paper is organized as follows. Section II presents a short literature review on current deep learning methods employed for COVID-19 diagnosis. Section III presents our proposed deep learning fusion architecture, while Section IV challenges our method against current techniques on CT and X-ray imagery. Experiments involve both two and four-class classification problems. Finally, Section V concludes this paper.

## II. Literature Review

The ongoing pandemic has initiated a massive research interest on deep learning-based COVID-19 diagnosis. This type of diagnosis mainly uses CT or X-ray medical images and can be distinguished in exploiting pre-trained or custom-designed models. It should be noted, that despite deep learning presents an overall appealing option, current methods use different evaluation datasets, and thus a direct comparison of the existing methods is not trivial. Therefore, in this section, we will not present the performance attained by each technique but a performance comparison will be presented only in the experimental Section III utilizing two common datasets. For better readability, in the following sub-sections we will present only a few representative techniques per model origin (pre-trained and custom), and data domain (CT and X-ray imagery). For further study on the advancements of deep learning strategies for COVID-19 diagnosis, the reader is referred to [21].

### A. Pre-trained models

This type of models extends the usability of existing deep networks trained in the visual domain, into classifying medical imagery for COVID-19 diagnosis after being properly re-trained using transfer learning. Several papers employ CT imagery, for example, Xu *et al*. [22] use the ResNet-18 model and Jin *et al*. the ResNet-152 [23], where the number indicates the longest convolutional – fully connected layer chain within the network. Wu *et al*. [24] propose a data-level fusion scheme where fused multi-view CT imagery is input to a ResNet-50 network [19]. Similar to other data domains, multi-view fusion attains higher classification than its single-view counterpart. Ardakani *et al*. [25] challenge the capability of current state-of-the-art deep networks on COVID-19 diagnosis. Their experiments involve AlexNet, VGG-16, VGG-19, SqueezeNet [26] GoogleNet, MobileNet-v2 [27], ResNet-18, ResNet-50, ResNet-101 and Xception [28]. Their work highlighted the very promising classification performance of deep learning, where ResNet-101 and Xception presented the highest accuracy.

X-ray imagery is also quite common for COVID-19 diagnosis. Apostolopoulos and Bessiana [29] challenge VGG-19, MobileNet-v2, Inception, Xception, and Inception-ResNet-v2 and attain an accuracy exceeding 96% on a three-class classification problem (COVID-19, bacterial and viral pneumonia, and normal). Loey *et al*. [30] increase the number of training images by employing a Generative Adversarial Network (GAN) [31]. The augmented training imagery is then used to perform transfer learning on AlexNet, GoogleNet, and ResNet-18. Experiments on a four-class classification problem (COVID-19, normal, pneumonia bacterial, pneumonia virus) highlight the great contribution of GAN to high classification accuracy.

### B. Custom models

Regardless of the data domain, the majority of the custom models are inspired by a pre-trained model. Considering CT imagery, Wang *et al*. [32] modify the inception concept [20] to present a smaller feature dimension. Accordingly, Liu *et al*. [33] alter DenseNet-264 [34] to present four dense blocks. Custom deep learning models also utilize X-ray imagery. Thus, Ozturk *et al*. [35] reduce the longest convolutional-fully connected layer chain of DarkNet [36] down to 17. Rahimzadeh and Attar [37] concatenate the deepest fully connected layers of Xception and ResNet-50 v2, and the concatenated feature vector is then input to a shallow convolutional neural network for classification. Li *et al*. [38] use discriminative cost-sensitive learning by combining fine-grained classification and cost-sensitive learning. Khobahi *et al*. [39] introduce CoroNet, a semi-supervised deep learning architecture based on auto-encoders.

## III. Deep Network Fusion Architecture

The suggested deep learning architecture innovatively fuses the inner layers of two pre-trained deep learning structures to interlay the inner structures of the deep networks and augment their strengths. Our proposed deep learning strategy opposes to simplistic decision-level approaches where the classification outcome of each network is incorporated in a decision function. Additionally, due to the backpropagation process during the training process, our method cannot be classified as a feature-fusion method but rather as a structure-fusion type of architecture.

Formally, we consider the layer-wise operation defined as,

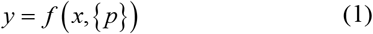

with *x* and *y* the input and the output vectors of the layers involved, and function *f* the operation applied using parameters. Thus, the deep network fusion of sub-network *a* at layer *i* with sub-network *b* at layer *j* is defined as,

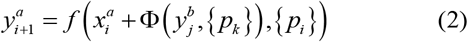

where Φ is the function linking the layers involved, which has parameters that highly depend on the cardinality relationship of 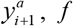, and 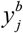. In general, we consider three distinct network fusion cases.

### A. Fusing fully connected layers

This case considers fusing two fully connected layers of each sub-network into a single layer. Given that the pre-trained sub-networks are originally trained on the same dataset, this fusion case involves Φ as an identity mapping process.

### B. Fusing layers of the same 2-dimensional cardinality

A common fusion requirement involves linking layers that contain 3-dimensional tensors presenting the same 2-dimensional cardinality over the first two dimensions and a different cardinality over the third one. In this case, function Φ cannot be a simple identity mapping process, but should linearly project 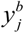 into the tensor size of 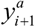. Thus, we define Φ as a 3-dimensional tensor,

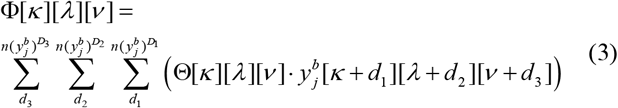

where *κ,λ,v,d*_*1*_,*d*_*2*_,*d*_*3*_ are the tensor coordinates, *n*(·)^*D*^ cardinality of a tensor over dimension D, and Θ a 3-dimensional kernel with elements originating from a Glorot initializer [40] with

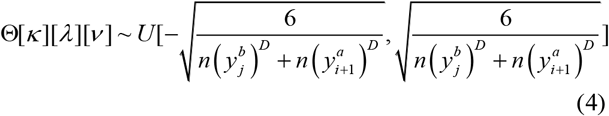

### C. Fusing layers of different cardinality

Layers of different depths within the same network or layers belonging to different networks are mainly of different cardinality 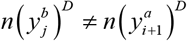, and thus fusing such layers requires extra care. Hence, for this instance, we consider a two-stage process, where initially function Φ is defined as in Eq. (3) but then is linearly interpolated in the 2-dimensional space for cardinality adjustment.

### D. Proposed network fusion architectures

Despite the suggested deep network fusion scheme can be applied to any deep network, in this work we utilize the GoogleNet [20] and ResNet-18 [19] deep networks. Both networks are pre-trained on ImageNet [14] involving 1.2 million images for training, 50,000 for validation, and 100,000 images for testing spreading over 1,000 object classes. GoogleNet is a 22-layered network with its major novelty being the inception module. The latter parallelizes the convolutional layers of different filter sizes with the max-pooling layer, aiming at better handling objects at multiple scales (Fig. 1 (a)). ResNet-18 is an 18-layered deep network that incorporates identity shortcut connections between the convolutional layers, converting a plain network into a residual (Fig. 1 (b)). The major advantage of residual networks is solving the vanishing gradient problem, which occurs when the deep gradients, from where the loss function is calculated, become zero during training.

**Fig. 1.**
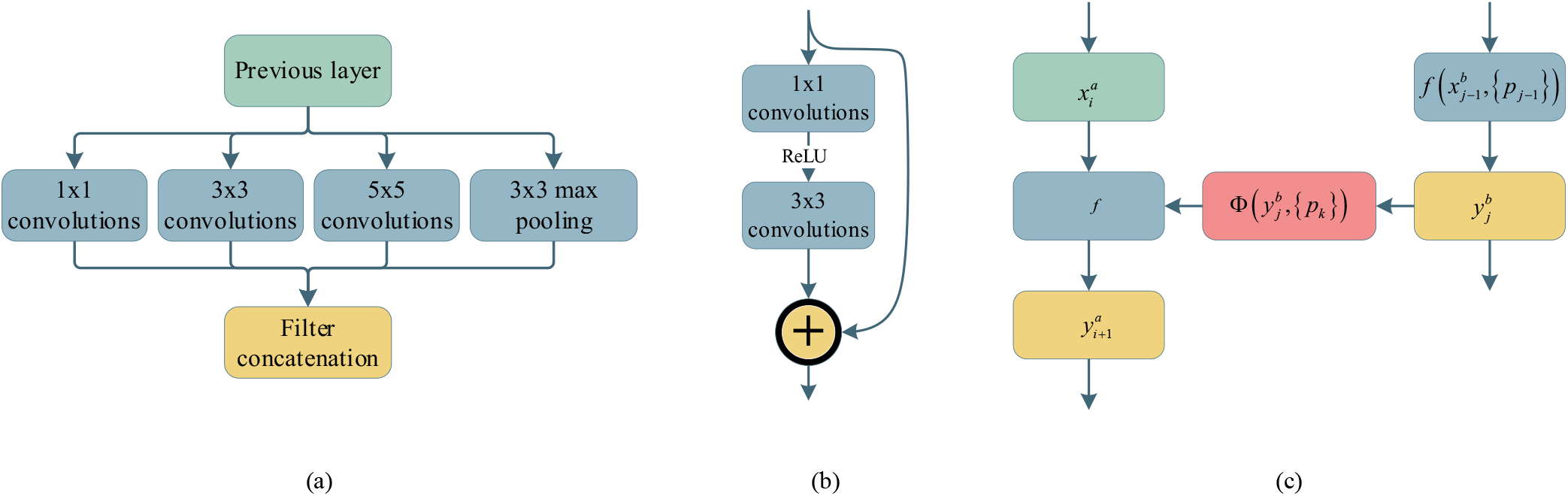
Generic representations of (a) GoogleNet inception layer (b) ResNet residual network (c) proposed deep network fusion

Thus, spurred by the powerful modules of GoogleNet and ResNet-18 we propose a layer fusion strategy (Fig. 1 (c)) linking several layers of these networks. Layer fusion may be performed between any layers of GoogleNet and ResNet by properly remapping the layer’s output to meet the input’s constraints, as per Eq. (2). However, for better readability, we constrain our fusion strategy to four cases.

a. *Case A*: This case considers fusing the deepest fully connected layer of each sub-network into a single layer. This fusion process is presented in Section III-A.
b. *Case B*: This instance extends *case A* and considers an additional fusion process between the *inception_3a* and the *residual_3a* layers of the GoogleNet and the ResNet-18, respectively. Opposing to *case A*, these layers present a dimensionality difference and thus Φ is estimated based on the strategy presented in Section III-B. The fused network is presented in Fig. 2.
c. *Case C*: This fusion strategy extends *case B* by additionally fusing the *inception_3b* and the *residual_3b* layers of the GoogleNet and the ResNet-18, respectively. For this case, Φ is estimated utilizing Eq. (3).
d. *Case D*: For this instance, we consider *case A* along with fusing the *inception_3a* and the *residual_4a* layers of the GoogleNet and the ResNet-18, respectively. Given the large dimensionality difference between the corresponding layers, Φ is estimated based on the technique presented in Section III-C such as to bridge the dimensionality gap between the *inception_3a* and the *residual_4a* layers.

**Fig. 2.**
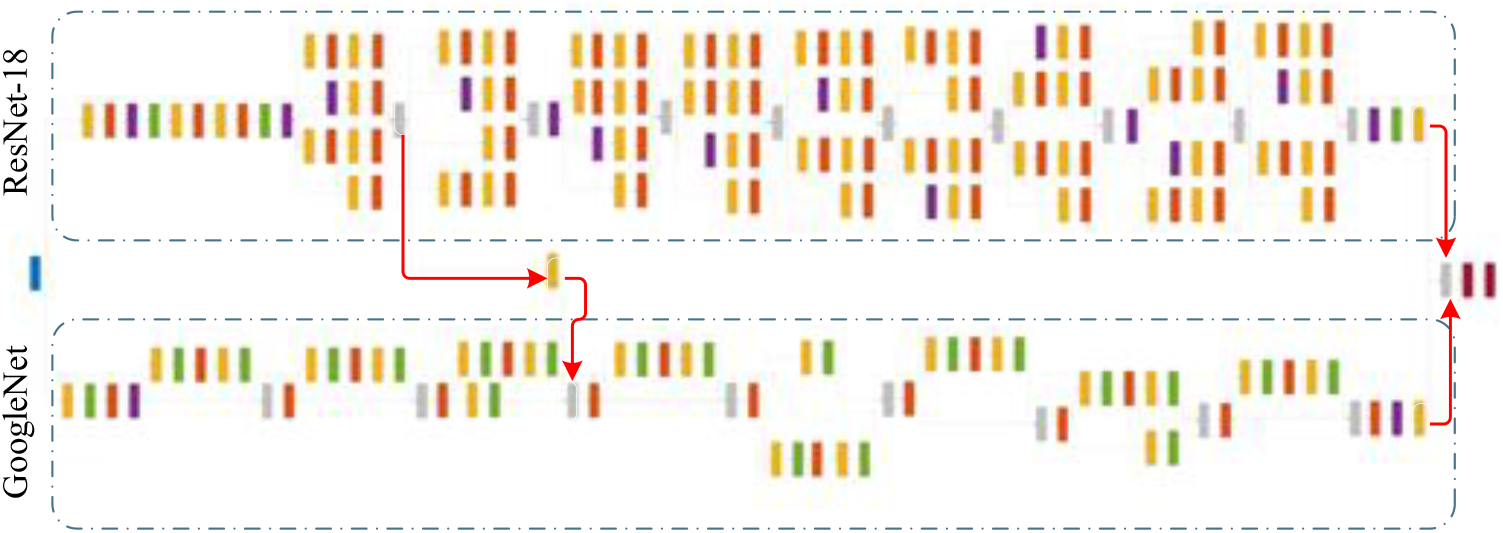
Deep network fusion architecture (*case B*) with red arrows highlighting the fused layers (building box layers, blue for input image, yellow for convolution and fully connected layers, red for activation layers, green for dropout, purple for pooling, gray for layer manipulation, red for output layers)

It should be noted that our deep learning fusion strategy contrasts [37] because in the latter work the authors do not alter the involved networks (Xception and ResNet-50 v2). In their work, they concatenate the features of the deepest fully connected layers and input the concatenated feature vector to a shallow convolutional neural network for classification. As a reminder, our strategy directly fuses the inner layers of the involved deep networks offering comprehensive deep learning fusion of the involving sub-networks.

It is worth noting that we also investigated fusing even more layers of various depths belonging to GoogleNet and ResNet-18. However, as presented in the experimental Section IV, the CT and X-ray imagery used for COVID-19 classification combined with the number of classification classes (two or four) produced an overall complexity that defined the required fusion complexity. Thus, increasing the fusion complexity between the two sub-networks did not improve performance, and thus these cases were omitted from this work. Finally, we also fused other state-of-the-art deep networks, i.e. ResNet-101, or even fused three deep sub-networks, but as already stated, the complexity of the problem investigated here did not require more complex fusion schemes than the ones presented in this paper.

## IV. Experiments

### A. Experimental setup

We validate the performance of the proposed deep network fusion scheme on two datasets of different data modalities. The first trial involves the Computed Tomography (CT) dataset of [41]. This is a two-class dataset containing CT scans from real patients in hospitals in Brazil and specifically, 1252 CT scans are positive for SARS-CoV-2 infection (COVID-19) and 1230 CT scans for patients non-infected by SARS-CoV-2. We also challenge our dataset on the X-ray dataset of [42] that is assembled based on the datasets of [43] and [44]. This is a four-class dataset, i.e. COVID-19, normal, pneumonia bacterial, and pneumonia virus, where each class contains 69, 79, 79, and 79 X-rays, respectively. For this dataset, the training to testing ratio is 6:1 for the COVID-19 class and 7:1 for the remaining classes. The reasoning for involving these datasets is to challenge our deep network fusion scheme under different data modalities and a small number of ground-truth imagery. Examples of both datasets are presented in Fig. 3.

**Fig. 3.**
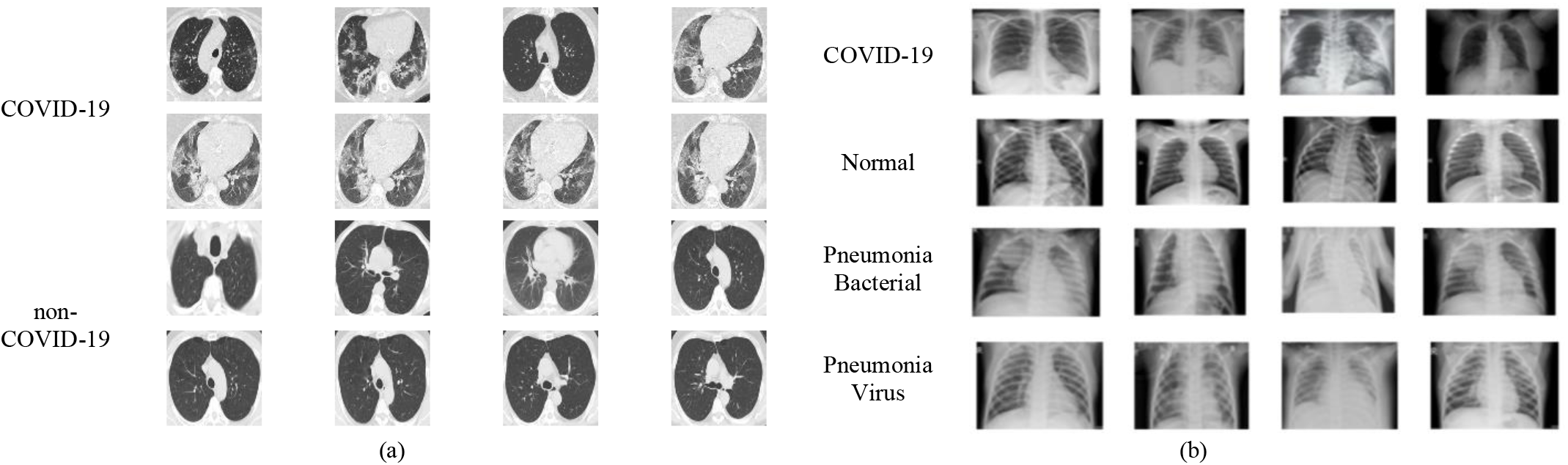
Datasets employed (a) two-class CT (b) four-class X-ray

We challenge our deep fused network utilizing four variants, i.e. *case A-D* as presented in Section III-C and exploiting the training parameters of Table I. Since both sub-networks utilized, i.e. GoogleNet and ResNet-18, are pre-trained, we apply on our suggested fused network the transfer learning technique [15] to fully exploit the classification capabilities of the initial networks. As expected, transfer learning is focusing on the fusion layers, where we set a learning rate for the weights and bias of 10. Additionally, we also apply data augmentation to reduce overfitting. For the latter, we employ a rather basic but still efficient approach that involves image rotation, translation, and shear. The data augmentation parameters are presented in Table I. All experiments are performed on an intel-i7 utilizing 24GB of RAM and an Nvidia GTX 1080Ti GPU.

**TABLE I.** Deep Network Fusion Training Parameters

| parameter | value |
| --- | --- |
| training |  |
| solver | sgdm |
| mini-batch size | 10 |
| initial training rate | $3 \cdot 10^{-4}$ |
| max epochs | 40 |
| learning rate | 10 |
| shuffle | every epoch |
| training : validation ratio | 7:3 |
| data augmentation |  |
| reflection | x-axis, y-axis |
| translation | x-axis, y-axis [-30,30] pixels |
| scale | [0.9,1.1] |
| shear | x-axis, y-axis [0.9,1.1]scale |

We evaluate the performance of our method against the competitor techniques of Table II using the accuracy and the F1-score metric defined as,

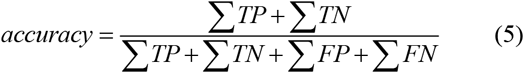

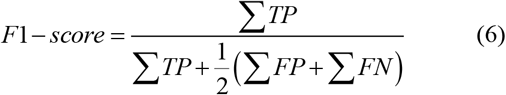

where for the True Positive (TP) case, the algorithm provides the hypothesis that the input image belongs to the COVID-19 class, which is correct. False Positive (FP), the algorithm provides a hypothesis that the input image belongs to the COVID-19 class, which is wrong. True Negative (TN), the algorithm classifies the input image as non-COVID-19 class, which is correct. False Negative (FN), the algorithm classifies the input image as a non-COVID-19 class, which is wrong. The definitions of TP, FP, TN, and FN presented above can be extended for the four-class classification problem considering a micro-averaged scheme per class.

**TABLE II.** Competitor Deep Network Setups

| Network | depth | parameters (millions) |
| --- | --- | --- |
| GoogleNet | 22 | 7 |
| ResNet-18 | 18 | 11.7 |
| AlexNet | 8 | 62.3 |
| VGG-16 | 16 | 138 |
| xDNN [41] | 16 | not defined |
| Inception v3 | 48 | 23.9 |
| ResNet-50 | 50 | 25.6 |
| DarkNet53 | 53 | 41.6 |
| Case A fusion | 40 | 12.4 |
| Case B fusion | 41 | 12.5 |
| Case C fusion | 42 | 12.7 |
| Case D fusion | 41 | 12.5 |

During all trials our network fusion architecture is challenged against solely using GoogleNet and ResNet-18, aiming at revealing the performance improvement by fusing these networks. Additionally, we also compare our fused architecture against the classic CNNs AlexNet [45] and VGG-16 [18], and finally against the current state-of-the-art deep networks of similar depth to our proposed fused network. Given that our fused network has a depth of 40 up to 42 depending on the fusion case, the competitor networks are Inception v3 [46], ResNet-50 [19], and DarkNet53 [36]. Finally, depending on the dataset employed, the comparison includes the methods presented by the current literature that utilize the corresponding dataset. Table II presents the competitor methods along with their depth and the number of parameters they contain.

### B. CT dataset

The first trial considers the computerized tomography imagery and challenges our GoogleNet/ RestNet-18 fusion architecture against the methods presented in Table II and the technique of Soares *et al*. [41] that exploits the same CT dataset. Experimental results are presented in Table III, where it is evident that the proposed deep network fusion scheme manages higher accuracy and F1-score than the competitor methods. More importantly, Table III demonstrates that fusing two pre-trained networks affords a higher classification rate than solely utilizing the same networks. Indeed, our proposed fusion architecture attains for the *case A* fusion 99.53% accuracy, while GoogleNet 96.78% and ResNet-18 97.05%, respectively. As expected, deep networks that are shallower than ours and have fewer parameters present lower performance. However, it is worth noting that our *case A* fusion strategy manages to perform better than networks of similar or even larger depth such as the Inception v3 (98.26% accuracy), ResNet-50 (98.90% accuracy), and DarkNet53 (98.70%). Hence, it is important to highlight that our fusion strategy combines the advantages of its core deep networks, while minimizes the corresponding drawbacks. This is because the backpropagation process during transfer learning cross-tunes and fine-tunes the weights and biases of the sub-networks. Regarding the rest of the fusion cases, despite these being inferior to *case A*, these still present very appealing solutions as they manage an accuracy metric that is higher than the vast majority of the competitor methods. This performance reduction is linked to the complexity of the two-class classification problem, indicating that an increased network depth is not necessary for this type of classification. Accordingly, our fusion strategy *case A* attains the highest F1-score, with the rest of the fusion cases following closely.

**TABLE III.**
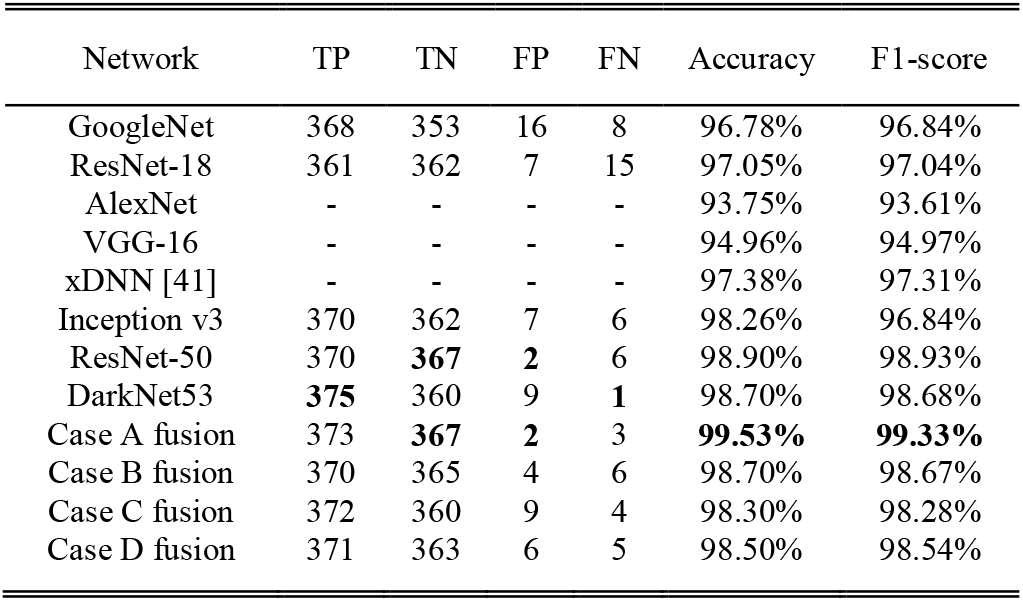
Two-class CT-based COVID-19 Diagnosis Performance (bold highlights top performance per metric)

| Network | TP | TN | FP | FN | Accuracy | F1-score |
| --- | --- | --- | --- | --- | --- | --- |
| GooleNet | 368 | 353 | 16 | 8 | 96.78% | 96.84% |
| ResNet-18 | 361 | 362 | 7 | 15 | 97.05% | 97.04% |
| AlexNet | - | - | - | - | 93.75% | 93.61% |
| VGG-16 | - | - | - | - | 94.96% | 94.97% |
| xDNN [41] | - | - | - | - | 97.38% | 97.31% |
| Inception v3 | 370 | 362 | 7 | 6 | 98.26% | 96.84% |
| ResNet-50 | 370 | **367** | **2** | 6 | 98.90% | 98.93% |
| DarkNet53 | **375** | 360 | 9 | **1** | 98.70% | 98.68% |
| Case A fusion | 373 | **367** | **2** | 3 | **99.53%** | **99.33%** |
| Case B fusion | 370 | 365 | 4 | 6 | 98.70% | 98.67% |
| Case C fusion | 372 | 360 | 9 | 4 | 98.30% | 98.28% |
| Case D fusion | 371 | 363 | 6 | 5 | 98.50% | 98.54% |

### C. X-ray dataset

Compared to the two-class CT dataset trials, this experiment is more challenging as it considers four classes and has fewer training samples. Similar to the experiments of Section IV-B, we compare our fusion strategy with the same mainstream deep networks, and also against the networks of [30], i.e. AlexNet, GooleNet, and ResNet-18 augmented via a GAN scheme.

Equally to the previous CT trials, fusing GoogleNet and ResNet-18 indeed enhances the classification accuracy. *Case A* fusion allows a 22.2% accuracy improvement over ResNet-18, the contributing sub-network that presents the highest accuracy among the two sub-networks used. Regarding GoogleNet, the performance gain of our fusion scheme is 25%. Interestingly, despite [30] combines ResNet-18 with a GAN data augmentation scheme, they achieve a lower accuracy compared to our basic data augmentation strategy. It is worth noting that compared to the two-class classification problem examined in Section III-B, four-class classification requires a higher fusion complexity. Specifically, from table IV it is evident that *case B* fusion manages 100% accuracy, while further increasing the fusion complexity (*case C* and *D*) presents a performance drop. This is because despite the four-class classification problem requires a more complex inter-layer fusion between the core sub-networks, extended complexity either in terms of linking more layers or linking deeper layers over fits the fused network and reduces its classification capability. Finally, our fusion strategy manages to present higher accuracy compared to deep networks of similar depth.

**TABLE IV.** Four-class x-ray-based COVID-19 Diagnosis Performance (bold highlights top performance per metric)

| Network | Accuracy |
| --- | --- |
| GooleNet | 75.00% |
| ResNet-18 | 77.80% |
| GAN/ AlexNet [30] | 66.67% |
| GAN/ GooleNet [30] | 80.56% |
| GAN/ ResNet-18 [30] | 69.46% |
| Inception v3 | 75.00% |
| ResNet-50 | 75.00% |
| DarkNet53 | 63.90% |
| Case A fusion | 80.60% |
| Case B fusion | **100%** |
| Case C fusion | 80.60% |
| Case D fusion | 75.00% |

## V. Conclusion

In this work, we present a novel deep learning fusion strategy that is appropriate for COVID-19 diagnosis. The proposed fusion technique is applied to current state-of-the-art pre-trained deep networks by interconnecting their internal layers. This is important as we fully exploit the knowledge of the core pre-trained deep learning models. Additionally, we create deep networks for enhanced classification performance without redesigning them. In this work we fuse GoogleNet and ResNet-18 and present four fusion cases at various layer depths and number of interconnections, presenting different model complexities.

We evaluate our fusion strategy on a two-class classification problem (COVID-19 vs. non-COVID-19) and a four-class problem (COVID-19, Normal, Pneumonia Bacterial and Pneumonia Virus) and demonstrate that our technique outperforms the core sub-networks of our fusion (GooleNet and ResNet-18) when these are solely used. Additionally, our evaluation demonstrates that the proposed method is more appealing compared to current state-of-the-art deep networks of similar complexity. Finally, we also demonstrate that the classification complexity (two vs. four-class classification) is linked to the fused network complexity.

Although we focus on COVID-19 diagnosis, our approach could be implemented to a great range of deep learning applications ranging from the medical and commercial domain to military and space applications.

## Data Availability

All data used are open-source

---

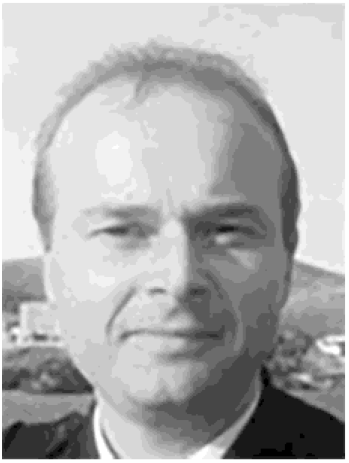

**Odysseas Kechagias-Stamatis** is currently a research Fellow in the Department of Electrical and Electronic Engineering at City University of London, UK. He received the MSc degree in Guided Weapon Systems and the Ph.D. degree in 3D ATR for missile platforms from Cranfield University, U.K. in 2011 and 2017 respectively. His research interests include visual odometry, 2D/3D object recognition and tracking, data fusion, and autonomy of systems.

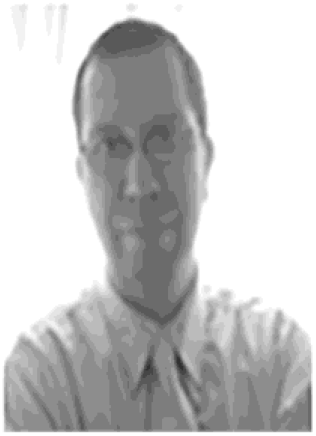

**Nabil Aouf** is currently a Professor of Robotics and Autonomous Systems in the Department of Electrical and Electronic Engineering at City University of London, UK. He has authored over 100 publications in high caliber in his domains of interest. His research interests are aerospace and defense systems, information fusion and vision systems, guidance and navigation, tracking, and control and autonomy of systems. He is an Associate Editor of the Imaging Science Journal

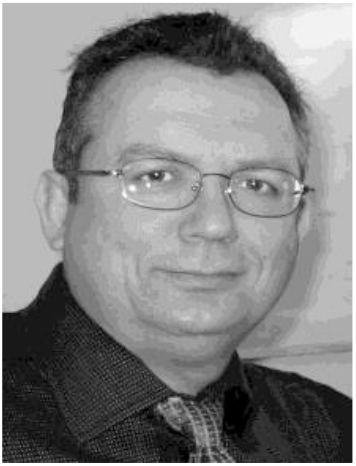

**John Koukos** is currently a Professor of Combat Systems, Naval Telecommunications & Electronics at the Hellenic Naval Academy, Piraeus, Greece. His research interests in the defense and aerospace fields include artificial intelligence, sensors, passive radar, military links and wireless networks, real time signal processing and applications for picosatellites and unmanned systems. Also GNSS applications and interference mitigation for unmanned systems.

